# Improving Retention and HIV Viral Suppression: A Cluster Randomised Pilot Trial of a Lay Counsellor Motivational Interviewing Training in South Africa

**DOI:** 10.1101/2024.09.20.24314048

**Authors:** Dorina Onoya, Tembeka Sineke, Idah Mokhele, Marnie Vujovic, Kate Holland, Robert A.C. Ruiter

## Abstract

We piloted a Motivational Interviewing (MI) training program for lay counsellors in South Africa (SA) to assess its impact on retention and viral suppression among newly diagnosed individuals living with HIV (PLHIV) at 12 months post-diagnosis. We randomized eight primary healthcare clinics (PHC) in Johannesburg to either the intervention clinics (n=4) where all lay counsellors were supported for 12 months before the PLHIV enrolment or the standard of care (n=4 clinics). Overall, 548 adults (≥ 18 years) PLHIV were recruited after HIV diagnosis from March 2020 to August 2021 (n=291 intervention, n=257 control). We conducted Poisson regression modelling to assess the intervention effect on patient attendance status (out of care – being ≥28 days late for the last appointment) and viral suppression (<50 copies/ml) at 12 months, reporting risk ratios (RR) with 95% confidence intervals (CIs). Of the 548 eligible participants enrolled, 56.3% were ≥28 days late (52.9% intervention vs 60.9% controls, RR 0.9, 95% CI: 0.7-1.0). Retention at 12 months positively correlated with baseline counselor MI skill levels (Cultivating change talk, RR 1.6, 95%CI: 1.1-2.5; Softening sustain talk, RR 1.5, 95%CI: 0.8-2.9; Empathy, RR 1.4, 95%CI: 1.0-1.8; Partnership RR 1.5, 95%CI: 1.2-1.9). Among those retained at 12 months, 65.3% of intervention participants were virally suppressed compared to 49.3% controls (RR 1.3, 95%CI: 1.0-1.7). Compared to control participants, the intervention group reported more positive counseling experiences, fewer concerns about HIV disclosure (RR 0.8 for high vs low-medium concerns, 95% CI: 0.7–1.0) and ART (RR 0.8, 95% CI: 0.7–1.0), and were more likely to express high confidence to take treatment in public (47.4% vs 28.8%, RR 1.4, 95% CI: 1.0–1.8) after counselling.

## INTRODUCTION

In 2015, UNAIDS introduced the ambitious 95-95-95 HIV program targets, ultimately aiming to achieve 86% viral suppression among people living with HIV (PLHIV) by 2030[1]. Despite concerted efforts, South Africa (SA) faces a significant challenge in reaching these goals[2, 3]. In 2021, out of 7.5 million people living with HIV, only 5.7 million had initiated antiretroviral therapy (ART)[4] and only 70% people living with HIV has achieved viral suppression[5]. To bridge this gap, a multifaceted approach focused on HIV testing, increased and sustained accessibility to ART, and improved retention in care is required, with additional emphasis on addressing stigma and discrimination, a major barrier to health-seeking behaviour and treatment adherence[6–8].

To bridge this gap, a multifaceted approach is required, focusing on HIV testing, increased and sustained accessibility to ART, and improved retention in care. Notably, addressing stigma and discrimination remains crucial, as these factors hinder health-seeking behaviour and treatment adherence [6–8]. In 2015, UNAIDS introduced the ambitious 95-95-95 HIV program targets, ultimately aiming to achieve 86% viral suppression among people living with HIV (PLHIV) by 2030[1]. Despite concerted efforts, South Africa (SA) faces a significant challenge in reaching these goals [2, 3]. In 2021, out of 7.5 million people living with HIV, only 5.7 million had initiated antiretroviral therapy (ART) [4] and only 70% people living with HIV had achieved viral suppression[5]. To bridge this gap, a multifaceted approach focused on HIV testing, increased and sustained accessibility to ART, and improved retention in care is required, with additional emphasis on addressing stigma and discrimination, a major barrier to health-seeking behaviour and treatment adherence [6–8].

Task-shifting has emerged as a pivotal strategy in the delivery of HIV services within primary healthcare (PHC) clinics, where lay counsellors undertake essential responsibilities such as HIV testing, counseling, and provision of psychosocial and support services [9, 10]. While the integration of lay counsellors has extended the reach of HIV care, lay counsellors often face constraints due to minimal training, which impacts their basic counseling skills, ultimately limiting their effectiveness [11, 12]. This limitation becomes particularly evident when balancing the demands of providing HIV testing services and delivering high-quality, patient-centered counseling. The current rapid ART initiation approaches necessitate a patient-centered and scalable ART demand creation strategy, allowing strategy [13–15]. This would allow early management of the complex interplay of sociocultural factors influencing health-seeking behaviors, ultimately establishing a pattern of ART and care adherence among PLHIV [16]. Lack of evidence-based standardised training that integrates patient-centered care approaches into routine healthcare practices, though, has resulted in limited awareness and uneven implementation [17]. In response to this imperative, we developed the Thusa-Thuso program, a Motivational Interviewing (MI) skills training and support program tailored for lay counselors[18],[19] MI is a client-centered, goal-oriented approach to communication and counseling that aims to elicit and strengthen an individual’s intrinsic motivation for positive behaviour change[20] The Thusa-Thuso training uses MI methods to equip counselors with necessary MI skills while also motivating them to commit to improving overall patient experience of quality counselling and prepare them for lifelong ART[18]. We showed that the Thusa-Thuso program improved counsellors’s MI adherent technical and relational skills progressively over the 12 months of support [21]. In this manuscript, we report the effect of the Thusa-Thuso counsellors training intervention on HIV patient ART retention and viral suppression by 12 months after HIV diagnosis.

## MATERIALS AND METHODS

### Study Design and Population

We conducted a cluster-randomized pilot trial (trial registration number: PACTR202212796722256) of newly diagnosed adult persons living with HIV (PLHIV) (≥18 years) enrolled from March 2020 to August 2021 across eight PHCs in Johannesburg (South Africa), randomly assigned to either the intervention condition (n=4 clinics), where all lay counsellors attended the Thusa-Thuso training and received support for 12 months before patient enrolment, or the standard of care support condition (n=4 clinics). Participant enrolment occurred simultaneously across randomization assignment randomisation assignments until the total sample size was achieved.

Trained study interviewers enrolled all participants through referrals from the testing counsellors at participating PHCs, immediately following post HIV-test counselling. Interviews with participants were conducted on the day of their enrolment into the study. PLHIV were eligible to participate if they self-reported that they were newly diagnosed. Patients with a documented history of prior ART use were excluded from the analytic dataset. Additionally, individuals who were either psychologically unable or too unwell to participate, unwilling to provide consent, or intending to seek treatment elsewhere were excluded from the study. Pregnant women at the time of HIV diagnosis were also excluded, as the initiation and care processes during pregnancy differ from those of non-pregnant women.

### Sample Size and Randomisation

We estimated that 616 participants (77 per site), would be needed to detect a 30% difference in the proportion of diagnosed patients retained on ART by 12 months between the intervention and control groups using an α of 0.05, 80% power, and a 1:1 allocation ratio. Additionally, we assumed an intra-cluster correlation of 0.01 and design effects of 1.8. Clinics were randomly assigned to either the intervention or control group with a 1:1 allocation, using a random number generator in Excel 2013 (Microsoft, Redmond, WA, USA). We randomly selected 20 clinics to assess the counsellor-level outcomes of the training intervention [21]. We used the CONSORT reporting guidelines [22]. For the PLHIV-level pilot study, a random selection of a subset of four intervention sites along with four control sites were selected using the same method.

### The Intervention: The Thusa-Thuso MI training program for lay counsellors

The Thusa-Thuso MI training program aimed to enhance general HIV counselling skills, cultivate sustained MI skills for patient-centered counselling, and ultimately improve ART uptake and retention in care among PLHIV. Lay counsellors underwent training in MI techniques through the Thusa-Thuso training program and received support for 12 months before patient enrolment. Further details about the program’s development and description can be found in the published reference [18].

The attainment of MI skills by lay counsellors was documented through Motivational Interviewing Treatment Integrity (MITI) coding of recorded counselling sessions during the 12 months of support before the patient trial.[21] MITI comprises two components: (1) global scores assigned on a five-point Likert scale characterising entire interactions as fostering change talks (CCT), softening sustain talk (SST), empathy (EM), or partnership (PA), and (2) behaviour counts that necessitate assessors to tally instances of specific counsellor behaviours, including complex reflections (CR), simple reflections (SR), affirmations, and emphasizing client autonom[23, 24]. Prior to patient enrolment, each counsellor was independently rated by two trained assessors, and the ratings for each session/participant were averaged to derive the final values [21]. We hypothesised that patients counselled and supported by MI-trained counsellors would achieve higher patient retention in care and HIV viral suppression outcomes 12 months after HIV diagnosis (the initial contact) compared to patients in control clinics.

### Data collection

Eligible and consenting PLHIV completed a structured baseline questionnaire on the day of HIV diagnosis. The questionnaire assessed baseline intention to initiate ART, and factors that could affect HIV treatment readiness including ART acceptability, socioeconomic status, HIV testing history, experiences with clinic services, alcohol use, HIV knowledge, and risk perceptions. The information sheet and questionnaire were available in English, Sesotho, and isiZulu, which are widely spoken and understood in Johannesburg. Participant follow-up occurred via medical record review from the date of HIV diagnosis to 12 months post-diagnosis or until the date of transfer/death in the first 12 months of care. Clinical information was collected from participants’ paper-based and electronic routine medical records, including laboratory test results.

### Analytic variables

The primary outcome, retention in care after 12 months, was defined as being within 28 days late for the last scheduled appointment in the first 12 months in care. During the study period, the vast majority of PLHIV received 28 treatment packs per visit. The secondary outcome was viral suppression which was defined as having a viral load of <50 copies/ml by 12 months in care. We also looked for counselling experience (pre-and post-test counselling) and preparedness for ART adherence (self-efficacy for adherence which was measured by and indication of how confident patients were to take medication in public, disclosure measured using a 16-item questionnaire with a 4-point scale (Strongly disagree) to four (Strongly agree) (Cronbach’s alpha = 0.89), and ART concerns measured using a 12-item questionnaire with a 4-point scale ranging from one (Strongly disagree) to four (Strongly agree) (Cronbach’s alpha = 0.83) and mean scores were categorized as low to medium and high ART concerns) and whether patients had received post-test counselling’s-demographic factors assessed include age, biological sex, highest education completed, English literacy, marital status, and the primary source of income.

### Data analysis

Effect on retention in care and viral suppression at 12 months after HIV diagnosis was assessed using Modified Poisson regression models, adjusting for baseline predictors and reporting Risk Ratios (RR) and 95% confidence intervals (95% CI). All statistical analyses were conducted in STATA 14™ (College Station, TX, USA).

### Ethical Review

Before any data collection, all participants provided written informed consent (administered by trained study staff before all data collection procedures in their preferred language). Confidentiality and anonymity were safeguarded by removing all identifiers, including both participant and facility names, from the analytic dataset. The study protocol was reviewed and approved by the Human Research Ethics Committee of the University of Witwatersrand (Wits HREC: M170579) and also by the Gauteng Provincial Department of Health (reference: GP_201711_028).

## RESULTS

### Characteristics of study participants

Out of 625 PLHIV screened, 592 (94.7%) met the eligibility criteria, with exclusions for reasons such as being underage, planning for treatment initiation elsewhere, pregnancy, prior HIV diagnosis or ART uptake, language barriers, and illness (Fig. 1). Ultimately, a sample of 548 enrolled participants was achieved, with 291 from intervention sites and 257 from control sites. The median age at HIV diagnosis was 34 years (Inter-quartile range (IQR): 29.0–41.0), and 61% of participants were female. Approximately 23% completed high school, 67.8% were proficient in English, and 64% were employed (Table 1). Medical records and visit history data at 12 months were available for review for only 449 participants (255 (87.3%) intervention and 192 (73.6%) controls).

**Figure 1.**
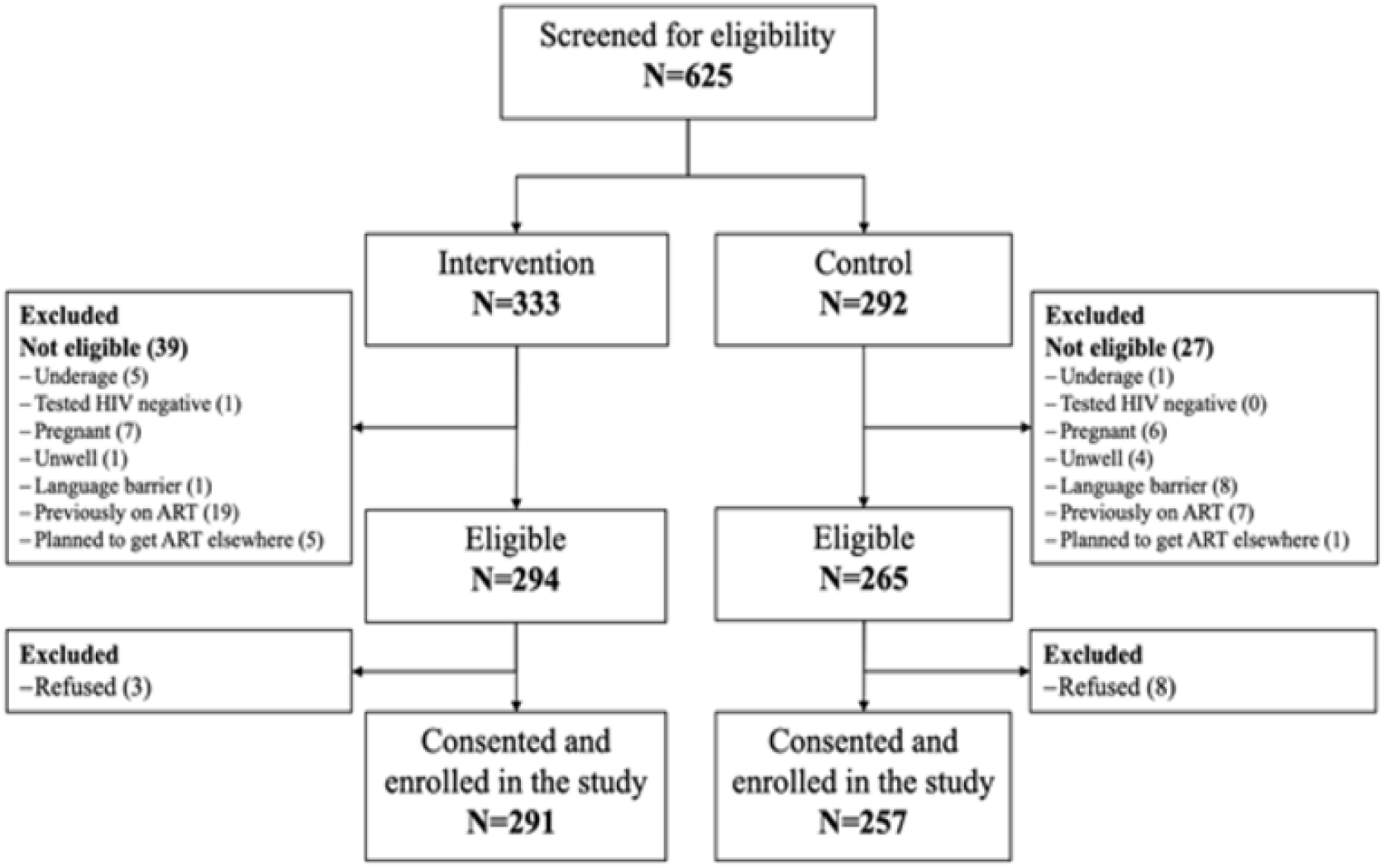
Recruitment, participant eligibility, and enrolment of newly diagnosed adults (≥18 years) at eight participating clinics in Johannesburg

**Table 1.** Characteristics of PLHIV by site randomisation allocation.

|  | Intervention |  | Control |  | Total |  |
| --- | --- | --- | --- | --- | --- | --- |
|  | n | Col (95% CI) | n | Col (95% CI) | n | Col (95% CI) |
| **Sex** |  |  |  |  |  |  |
| Female | 186 | 0.6 (0.6-0.7) | 149 | 0.6 (0.5-0.6) | 335 | 0.6 (0.6-0.7) |
| Male | 105 | 0.4 (0.3-0.4) | 108 | 0.4 (0.4-0.5) | 213 | 0.4 (0.3-0.4) |
| **Age (years)** |  |  |  |  |  |  |
| 18-29.99 | 79 | 0.3 (0.2-0.4) | 76 | 0.3 (0.2-0.4) | 155 | 0.3 (0.2-0.3) |
| 30-39.99 | 137 | 0.5 (0.4-0.6) | 108 | 0.4 (0.3-0.5) | 245 | 0.4 (0.4-0.5) |
| 40+ | 75 | 0.3 (0.2-0.3) | 73 | 0.3 (0.2-0.3) | 148 | 0.3 (0.2-0.3) |
| **Highest education level** |  |  |  |  |  |  |
| Primary school or less | 54 | 0.2 (0.1-0.3) | 25 | 0.1 (0.0-0.2) | 79 | 0.1 (0.1-0.2) |
| High school | 163 | 0.6 (0.4-0.7) | 115 | 0.5 (0.4-0.5) | 278 | 0.5 (0.4-0.6) |
| Completed Matric | 39 | 0.1 (0.1-0.2) | 88 | 0.3 (0.2-0.5) | 127 | 0.2 (0.1-0.4) |
| Post-matric | 31 | 0.1 (0.0-0.2) | 27 | 0.1 (0.0-0.3) | 58 | 0.1 (0.1-0.2) |
| **English proficiency** |  |  |  |  |  |  |
| I can read very well | 208 | 0.7 (0.7-0.8) | 162 | 0.6 (0.4-0.8) | 370 | 0.7 (0.6-0.8) |
| I can read somewhat | 55 | 0.2 (0.1-0.2) | 82 | 0.3 (0.2-0.5) | 137 | 0.3 (0.2-0.4) |
| I cannot read | 24 | 0.1 (0.1-0.1) | 13 | 0.1 (0.0-0.1) | 37 | 0.1 (0.0-0.1) |
| **Marital status** |  |  |  |  |  |  |
| Single, no partner | 58 | 0.2 (0.1-0.3) | 49 | 0.2 (0.1-0.3) | 107 | 0.2 (0.1-0.3) |
| In a relationship | 194 | 0.7 (0.6-0.8) | 177 | 0.7 (0.6-0.8) | 371 | 0.7 (0.6-0.8) |
| Married | 35 | 0.1 (0.1-0.2) | 31 | 0.1 (0.1-0.3) | 66 | 0.1 (0.1-0.2) |
| **Living with partner** |  |  |  |  |  |  |
| Yes | 133 | 0.5 (0.3-0.6) | 116 | 0.5 (0.4-0.5) | 249 | 0.5 (0.4-0.5) |
| No | 96 | 0.3 (0.2-0.5) | 91 | 0.4 (0.3-0.4) | 187 | 0.3 (0.3-0.4) |
| N/A -not in a relationship | 58 | 0.2 (0.1-0.3) | 49 | 0.2 (0.1-0.3) | 107 | 0.2 (0.1-0.3) |
| **Employment status** |  |  |  |  |  |  |
| Employed | 173 | 0.6 (0.5-0.7) | 174 | 0.7 (0.6-0.8) | 347 | 0.6 (0.6-0.7) |
| Unemployed | 114 | 0.4 (0.3-0.5) | 83 | 0.3 (0.2-0.4) | 197 | 0.4 (0.3-0.4) |
| **Income source** |  |  |  |  |  |  |
| Paid job, salary or business | 175 | 0.6 (0.5-0.7) | 175 | 0.7 (0.6-0.8) | 350 | 0.6 (0.6-0.7) |
| Partner/Family | 86 | 0.3 (0.2-0.4) | 70 | 0.3 (0.2-0.4) | 156 | 0.3 (0.2-0.4) |
| Other | 26 | 0.1 (0.1-0.1) | 11 | 0.0 (0.0-0.1) | 37 | 0.1 (0.0-0.1) |
| **Breadwinner of household** |  |  |  |  |  |  |
| Yes | 165 | 0.6 (0.5-0.7) | 152 | 0.6 (0.5-0.7) | 317 | 0.6 (0.5-0.7) |
| No | 122 | 0.4 (0.3-0.5) | 104 | 0.4 (0.3-0.5) | 226 | 0.4 (0.3-0.5) |
| **Location of primary house** |  |  |  |  |  |  |
| Current house | 126 | 0.4 (0.2-0.7) | 53 | 0.2 (0.1-0.3) | 179 | 0.3 (0.2-0.5) |
| Another province | 75 | 0.3 (0.1-0.5) | 104 | 0.4 (0.3-0.5) | 179 | 0.3 (0.2-0.5) |
| Another country | 81 | 0.3 (0.2-0.5) | 100 | 0.4 (0.3-0.5) | 181 | 0.3 (0.2-0.4) |
| **HIV knowledge** |  |  |  |  |  |  |
| Low HIV knowledge | 94 | 0.3 (0.2-0.5) | 74 | 0.3 (0.2-0.5) | 168 | 0.3 (0.2-0.4) |
| High HIV knowledge | 188 | 0.7 (0.5-0.8) | 178 | 0.7 (0.5-0.8) | 366 | 0.7 (0.6-0.8) |
| **Baseline CD4<350** |  |  |  |  |  |  |
| No | 72 | 0.4 (0.3-0.5) | 51 | 0.3 (0.3-0.4) | 123 | 0.4 (0.3-0.4) |
| Yes | 122 | 0.6 (0.5-0.7) | 101 | 0.7 (0.6-0.7) | 223 | 0.6 (0.6-0.7) |

### Visit attendance status at 12 months post-HIV diagnosis

Overall, 253/449 (56.3%) of the participants were 28 days late for their last scheduled appointment by 12 months in care, 136 (52.9%) in the intervention site versus 117 (60.9%) in the control sites (RR 0.9, 95% CI: 0.7-1.0) (Table 2). A lower proportion, 43.2%, could be classified as being 90 days late for their last appointment (41.2% for the intervention vs 45.8% for the controls, Fig 2). Within the intervention group, counsellors’ endpoint MI skill scores positively correlated with patient retention at 12 months in care and retention rates increased proportional to increasing MI skill levels (Fig 3). Within the intervention group, the proportion of PLHIV retained increased from 44% to 75% with a higher mean CCT score (CCT score 2.8 to 4) and increased from 44% to 54.6% with a higher mean SST score (SST score 2.5 to 3.7). Retention increased from 43.8% to 75% with a higher mean empathy score (Empathy score 2.7 to 4.7) and increased from 44.4% to 75% with a higher mean partnership score (3.0 to 4.5) (CCT, (crude Risk Ratio (cRR) RR 1.6, 95% CI: 1.1-2.5; SST, RR 1.5, 95% CI: 0.8-2.9; Empathy, cRR 1.4, 95% CI: 1.0-1.8; Partnership RR 1.5, 95% CI: 1.2-1.9).

**Figure 2.**
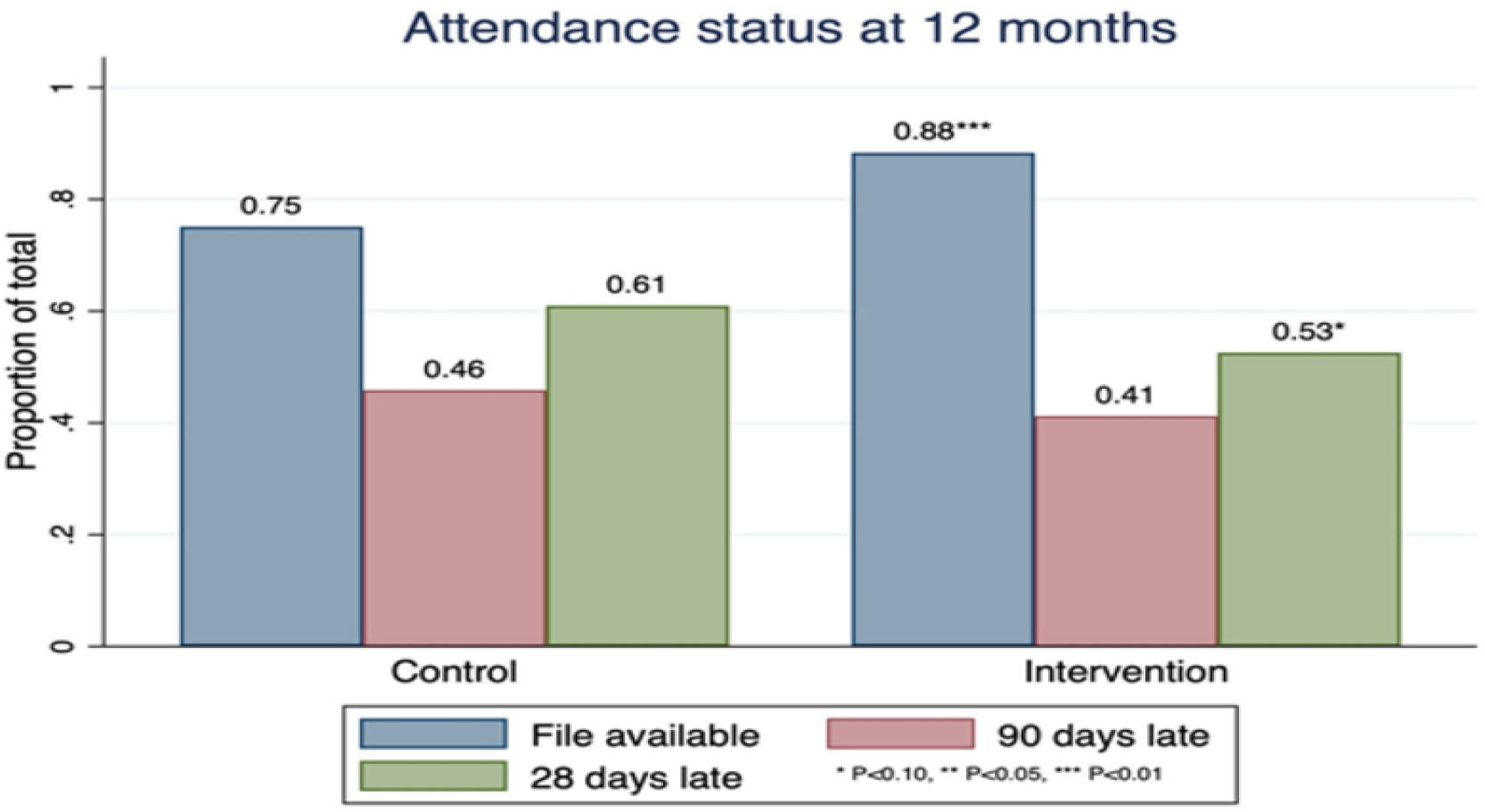
**Study participants’ clinic attendance status at 12 months post-HIV diagnosis for the intervention and control**

**Figure 3.**
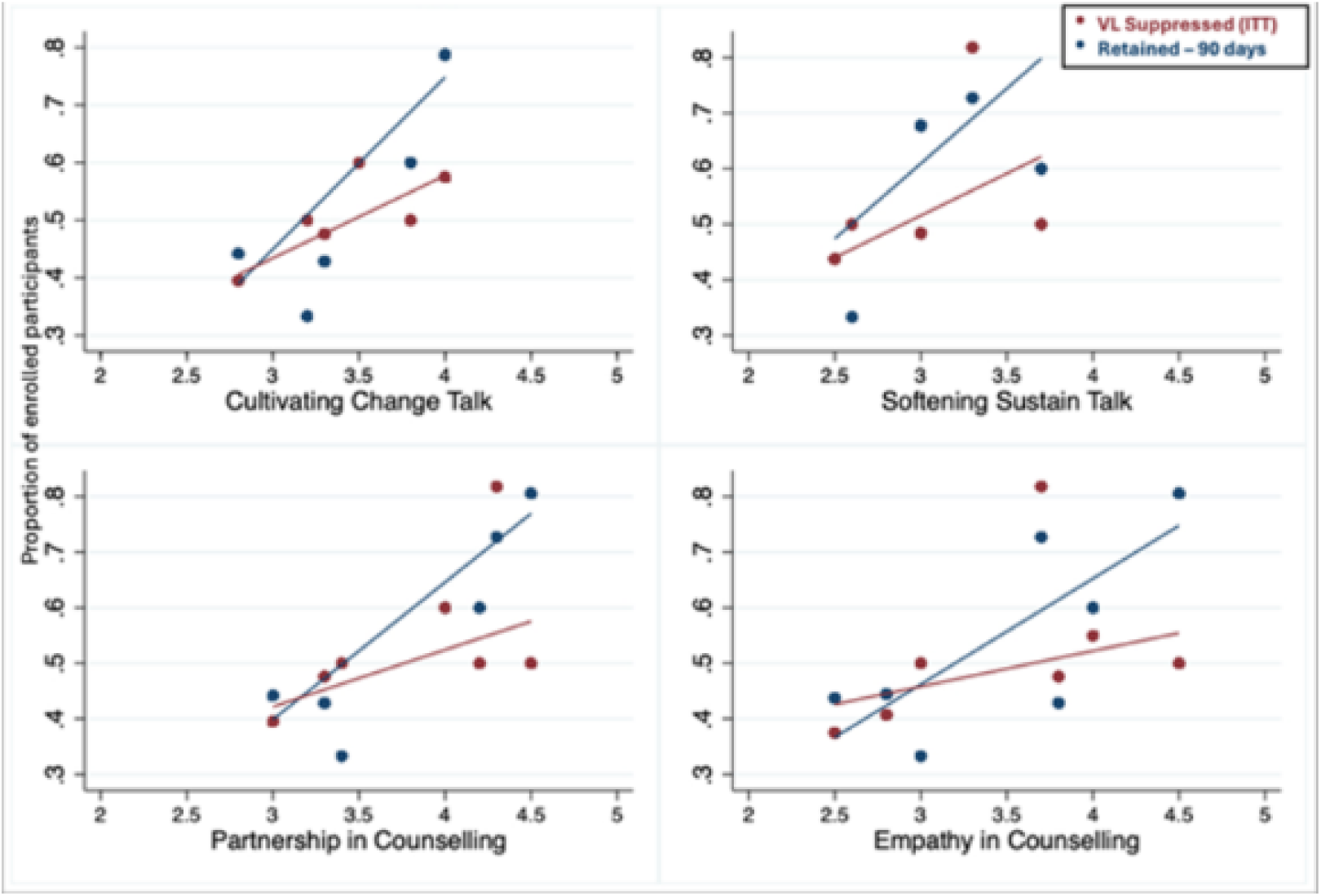
**Change in the proportion retained or VL suppressed at 12 months post-HIV diagnosis by counsellor MI skill level (SST, CCT, Empathy, Partnership)**

**Table 2.**
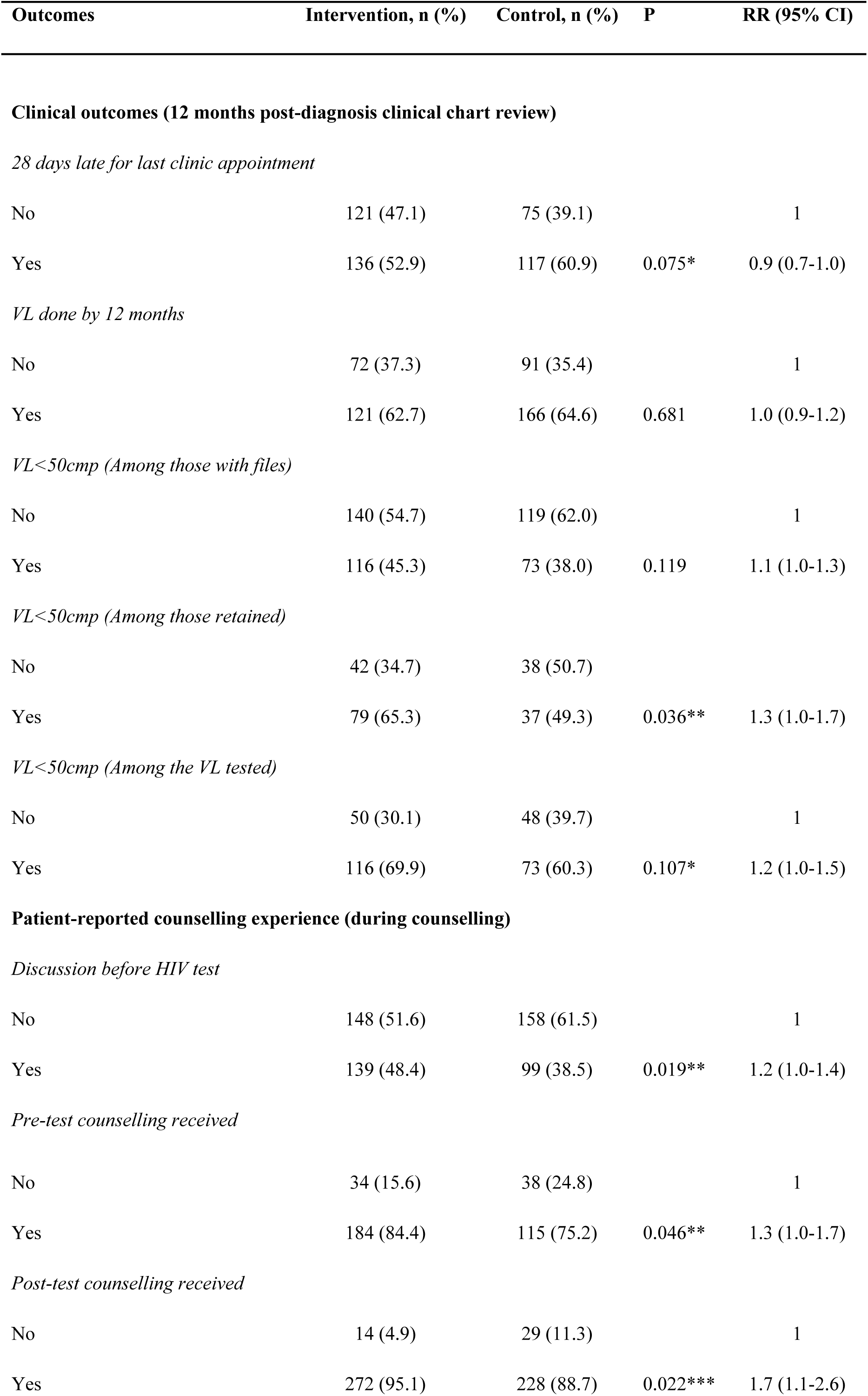
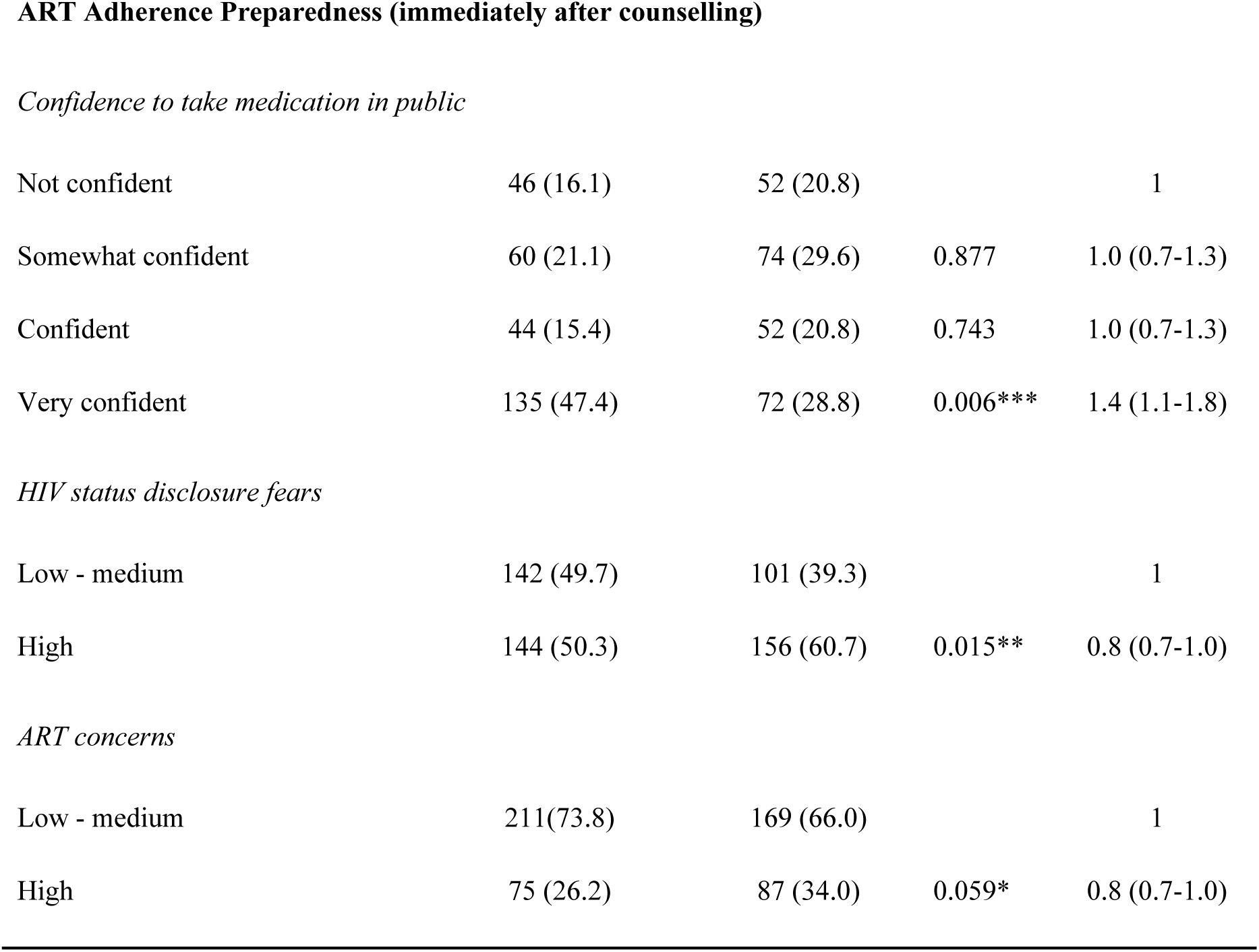
Intervention effect on engagement in care, ART adherence preparedness and patient-reported counselling experience.

| Outcomes | Intervention, n (%) | Control, n (%) | P | RR (95% CI) |
| --- | --- | --- | --- | --- |
| **Clinical outcomes (12 months post-diagnosis clinical chart review)***28 days late for last clinic appointment* |  |  |  |  |
| No | 121 (47.1) | 75 (39.1) |  | 1 |
| Yes | 136 (52.9) | 117 (60.9) | 0.075* | 0.9 (0.7-1.0) |
| *VL done by 12 months* |  |  |  |  |
| No | 72 (37.3) | 91 (35.4) |  | 1 |
| Yes | 121 (62.7) | 166 (64.6) | 0.681 | 1.0 (0.9-1.2) |
| *VL<50cmp (Among those with files)* |  |  |  |  |
| No | 140 (54.7) | 119 (62.0) |  | 1 |
| Yes | 116 (45.3) | 73 (38.0) | 0.119 | 1.1 (1.0-1.3) |
| *VL<50cmp (Among those retained)* |  |  |  |  |
| No | 42 (34.7) | 38 (50.7) |  |  |
| Yes | 79 (65.3) | 37 (49.3) | 0.036** | 1.3 (1.0-1.7) |
| *VL<50cmp (Among the VL tested)* |  |  |  |  |
| No | 50 (30.1) | 48 (39.7) |  | 1 |
| Yes | 116 (69.9) | 73 (60.3) | 0.107* | 1.2 (1.0-1.5) |
| **Patient-reported counselling experience (during counselling)***Discussion before HIV test* |  |  |  |  |
| No | 148 (51.6) | 158 (61.5) |  | 1 |
| Yes | 139 (48.4) | 99 (38.5) | 0.019** | 1.2 (1.0-1.4) |
| *Pre-test counselling received* |  |  |  |  |
| No | 34 (15.6) | 38 (24.8) |  | 1 |
| Yes | 184 (84.4) | 115 (75.2) | 0.046** | 1.3 (1.0-1.7) |
| *Post-test counselling received* |  |  |  |  |
| No | 14 (4.9) | 29 (11.3) |  | 1 |
| Yes | 272 (95.1) | 228 (88.7) | 0.022*** | 1.7 (1.1-2.6) |
| *Confidence to take medication in public* |  |  |  |  |
| Not confident | 46 (16.1) | 52 (20.8) |  | 1 |
| Somewhat confident | 60 (21.1) | 74 (29.6) | 0.877 | 1.0 (0.7-1.3) |
| Confident | 44 (15.4) | 52 (20.8) | 0.743 | 1.0 (0.7-1.3) |
| Very confident | 135 (47.4) | 72 (28.8) | 0.006*** | 1.4 (1.1-1.8) |
| *HIV status disclosure fears* |  |  |  |  |
| Low - medium | 142 (49.7) | 101 (39.3) |  | 1 |
| High | 144 (50.3) | 156 (60.7) | 0.015** | 0.8 (0.7-1.0) |
| *ART concerns* |  |  |  |  |
| Low - medium | 211(73.8) | 169 (66.0) |  | 1 |
| High | 75 (26.2) | 87 (34.0) | 0.059* | 0.8 (0.7-1.0) |

### VL suppressed at 12 months

Among those with available medical records (Table 2), 189/449 (42.2%) of the participants were virally suppressed (VL <50 copies/ml), 116 (45.3%) in the intervention site versus 73 (38.0%) in the control sites (RR 1.1, 95%CI: 1.0-1.3). Among PLHIV who were retained in care (within 28 days of appointment by 12 months), 65.3% of the intervention site participants were virally suppressed versus 49.3% in the control sites (RR 1.3, 95% CI: 1.0-1.7). Among those who were tested (Figure 4), 69.9% of the intervention participants vs 60.3% of control participants were virally suppressed (RR 1.2, 95%CI: 1.0-1.5). Similarly, to the retention outcome (within the intervention group, increasing counsellor MI skills resulted in modestly higher PLHIV VL suppression at 12 months (Fig 2.) (CCT, cRR 1.2, 95% CI: 1.2-1.3; SST, RR 1.3, 95% CI: 0.8-2.2; Empathy, RR 1.1, 95% CI: 0.9-1.4; Partnership, RR 1.2, 95%CI: 1.1-1.3).

**Figure 4.**
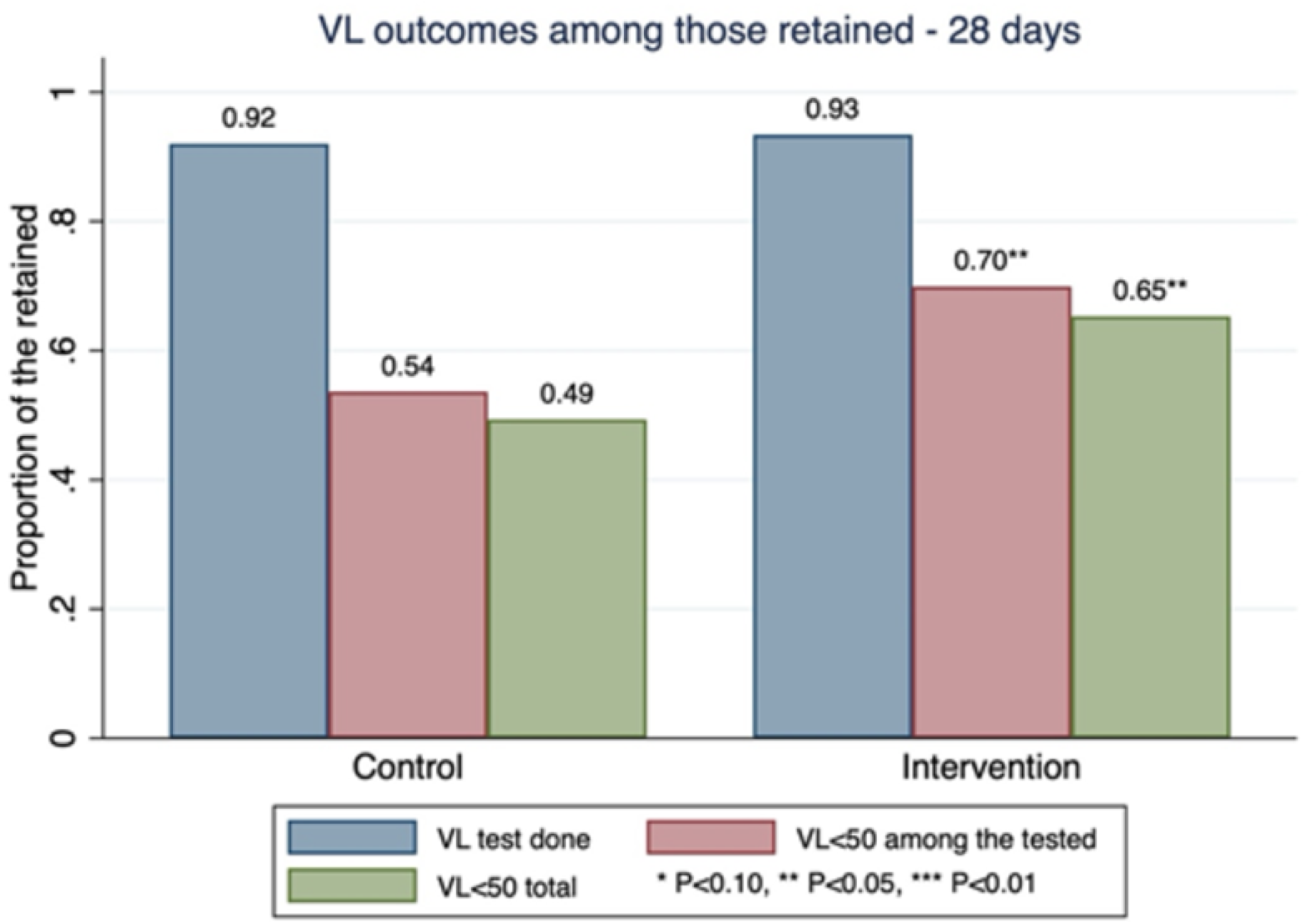
**Study participants’ VL status at 12 months post-HIV diagnosis for the intervention and control clinics**

### Counselling Experience and ART Adherence Preparedness

We compared intervention and control PLHIV participants on immediate outcomes of HIV counselling. Compared to control sites, participants at intervention sites were more likely to indicate having had a discussion with the counsellor pre-test (48.4% vs 38.5%, RR 1.2, 95%CI: 1.0-1.4) and also reported receiving pre-test counselling ( 84.4% vs 75.2% for the intervention and control sites, respectively, RR 1.3, 95%CI: 1.0-1.7) and post-test counselling (95.1% vs 88.7% for the intervention and control sites, respectively, RR 1.7, 95% CI: 1.1–2.6).

Additionally, compared to controls, intervention participants were more likely to express high confidence to take treatment in public (Fig 5) (47.4% vs 28.8%, RR 1.4, 95% CI: 1.0–1.8) after counselling. Furthermore, intervention participants were less likely to report HIV disclosure concerns (50.4% intervention vs 60.7% controls, RR 0.8, 95% CI: 0.7–1.0), and less likely to report high concerns about ART (45.7% intervention vs 54.3% control, RR 0.8 for high vs low-medium concerns, 95% CI: 0.7–1.0) than controls.

**Figure 5.**
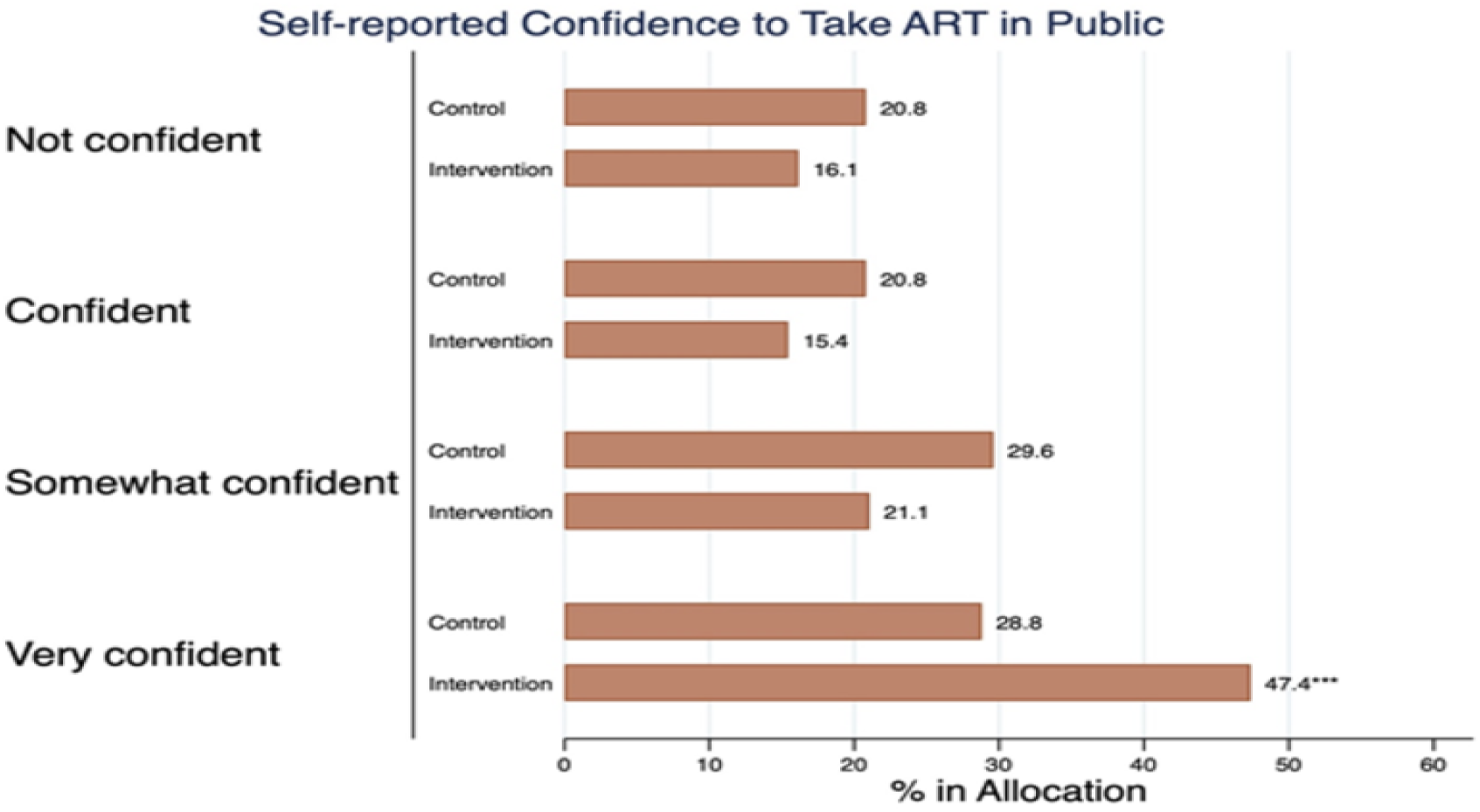
**Study participants’ confidence to take ART in public immediately after counselling for the intervention and control clinics**

## DISCUSSION

This cluster randomized pilot trial of a previously described MI training program for lay counselors [21] aimed to assess the impact of trained counselors on outcomes for newly diagnosed PLHIV. Overall, a significant proportion of study participants were out of care at 12 months, indicating challenges in maintaining consistent engagement during the COVID-19 pandemic period when the study was conducted, a phenomenon that was generally observed across primary-care settings in South African [25, 26].

However, positive correlations between counselor MI skills and patient retention at 12 months underscore the potential impact of MI counseling on long-term engagement, consistent with previous findings demonstrating the efficacy of MI counseling in enhancing adherence to ART and promoting better health outcomes among individuals living with HIV/AIDS[27, 28]. Our study found that increasing MI skill levels, particularly in communication, support, empathy, and partnership, were associated with improved retention rates, emphasizing the importance of patient-centered care in HIV management. Other studies, particularly among those with chronic conditions, have also shown that clients are more likely to stay engaged in interventions or treatment programs when they perceive their practitioners as empathetic, supportive, and collaborative partners in the change process [29]. In a similar setting, MI counselling was shown to reduce delays in early infant diagnosis testing schedules for HIV-exposed infants, further showing the potential of MI as an effective intervention in the context of HIV management [30]. Together, these findings contribute to the growing body of evidence supporting MI’s role in enhancing adherence to healthcare protocols and promoting positive outcomes for clients across a range of settings and populations [18].

We also observed higher rates of viral load suppression in the intervention sites compared to control sites, with increasing counselor MI skills associated with higher viral load suppression rates within the intervention group, particularly among those retained in care by 12 months. These findings are consistent with another study which showed that a participants’ intrinsic motivation plays a pivotal role in enhancing screening uptake and therapy adherence [31, 32].

In addition to clinical outcomes, participants in the intervention group reported more positive counseling experiences, lower disclosure fears, and higher confidence in taking treatment in public than the controls. These measures indicate higher ART adherence readiness in the intervention group which is an important outcome of HIV counselling and MI support. MI is designed to empower individuals to feel confident in their choices and their self-efficacy for attaining health and wellness goals [33]. Addressing HIV disclosure and ART concerns is particularly important since disclosure concerns are consistently shown to be a major barrier in HIV care [34, 35].

### Study limitation

Several limitations of the study warrant consideration. The sample predominantly comprised clinics and PLHIV from the City of Johannesburg in the Gauteng province, potentially limiting the generalizability of findings to broader settings and populations. Reliance on self-reported measures for counseling experiences, confidence levels, and concerns about ART may have introduced social desirability bias. Moreover, variability in counselor skill levels within intervention sites could have influenced intervention effectiveness and contributed to outcome heterogeneity. Conducting the study during the COVID-19 pandemic and associated lockdown measures may have influenced participant access to healthcare services and overall study outcomes. Additionally, incomplete medical records and visit history data at the 12-month mark may have introduced selection bias. Lastly, as a pilot cluster trial involving only a subset of primary healthcare facilities in the Gauteng province, the study design may not fully capture the complexities of implementing MI training for lay counselors in real-world settings. Larger-scale randomized controlled trials with longer follow-up periods and diverse participant populations are necessary to validate intervention effectiveness and sustainability.

## CONCLUSION

In conclusion, the trial suggests that MI training for lay counselors can positively impact both retention in care and viral load suppression among PLHIV. The findings highlight the importance of counselor skills, communication, and addressing patient concerns in achieving favorable HIV-related outcomes. Further research and interventions should explore ways to enhance counseling strategies and address contextual factors influencing retention and viral suppression.

## Data Availability

The datasets generated and/or analyzed during the current study are available from the Health Economics and Epidemiology Research Office for researchers who meet the criteria for access to confidential data and with permission from the owners of the data. Contact the organization at for additional information regarding data access.

## Acknowledgments

The authors wish to thank the all study participants and staff at all PHC facilities involved in the study. Additionally, they would like to acknowledge their counseling and data collection team’s commitment and tireless efforts: Alice Kono, Sinetemba Madlala, Nonhlanhla Tshabalala, Pertunia Manganye, Phuthi Moshupja, Michael Mothapo, Simangele Sigasa, and Zanele Walaza.

## Contributors

DO conceptualised the project. IM and TS were involved in the study implementation. DO and TS analysed the data and wrote the original draft manuscript. IM, KH, MV and RR provided feedback on the manuscript. All authors assisted in interpreting the results and critically reviewed and approved the final version of the manuscript.

## Funding

This study has been made possible by the generous support of the American People and the President’s Emergency Plan for AIDS Relief (PEPFAR) through United States Agency for International Development (USAID) under the terms of Cooperative Agreement 72067419CA00004 to the Wits Health Consortium. DO, TS and KS were also supported by U.S. National Institutes of Health 1R01AI152149. The contents are the responsibility of the authors and do not necessarily reflect the views of PEPFAR, USAID, NIH or the United States Government.

## Declaration of interests

The authors confirm that the research was performed in accordance with the Declaration of Helsinki. The study was approved by the Human Research Ethics Committee (Medical) of the University of the Witwatersrand (Wits HREC M170579). All participants provided written informed consent to participate in the study. Informed consent was administered in the participant’s preferred language (English, Sotho or Zulu). All personal identifiers, including participants’ and facility names, were removed from the final analytic dataset.

## Notes

### Competing Interest Statement

The authors have declared no competing interest.

### Clinical Trial

PACTR202212796722256

### Author Declarations

The study protocol was reviewed and approved by the Human Research Ethics Committee of the University of Witwatersrand (Wits HREC: M170579) and also by the Gauteng Provincial Department of Health (reference: GP_201711_028).

## REFERENCES

1. Joint United Nations Programme on HIV/AIDS (UNAIDS). Understanding fast-track: Accelerating action to end the AIDS epidemic by 2030. UNAIDS Switzerland; 2015.

2. NDoH. National Policy on HIV Pre-exposure Prophylaxis (PrEP) and Test and Treat (T&T). Pretoria: National Department of Health. 2016.

3. NDoH. Fast tracking implementation of the 90-90-90 strategy for HIV, through implementation of the test and treat (TT) policy and same-day anti-retroviral therapy (ART) initiation for positive patients. Pretoria, South Africa: National Department of Health. 2017.

4. Joint United Nations Programme on HIV/AIDS (UNAIDS). Country factsheets: SOUTH AFRICA 2021. UNAIDS data 2020. Available from: https://www.unaids.org/en/regionscountries/countries/southafrica.

5. NDoH. Annual report 2021/2022. Pretoria, South Africa, 2022.

6. Chan BT, Tsai AC. HIV stigma trends in the general population during antiretroviral treatment expansion: analysis of 31 countries in sub-Saharan Africa, 2003–2013. JAIDS Journal of Acquired Immune Deficiency Syndromes. 2016;72(5):558–64.

7. Horter S, Bernays S, Thabede Z, Dlamini V, Kerschberger B, Pasipamire M, et al. “I don’t want them to know”: how stigma creates dilemmas for engagement with treat-all HIV care for people living with HIV in Eswatini. African journal of AIDS research. 2019;18(1):27–37.

8. Kalichman SC, Mathews C, Banas E, Kalichman MO. Treatment adherence in HIV stigmatized environments in South Africa: stigma avoidance and medication management. International journal of STD & AIDS. 2019;30(4):362–70.

9. Dewing S, Mathews C, Cloete A, Schaay N, Simbayi L, Louw J. Lay counselors’ ability to deliver counseling for behavior change. Journal of consulting and clinical psychology. 2014;82(1):19.

10. Organization WH. Task shifting: rational redistribution of tasks among health workforce teams: global recommendations and guidelines. 2007.

11. Lewin S, Dick J, Pond P, Zwarenstein M, Aja GN, van Wyk BE, et al. Lay health workers in primary and community health care. Cochrane database of systematic reviews. 2005;(1).

12. Mokhele I, Sineke T, Vujovic M, Ruiter RAC, Onoya D. Who is providing HIV testing services? The profile of lay counsellors providing HIV testing services in Johannesburg, South Africa in the treat-all era. BMC Health Services Research. 2023;23(1):1372. doi: 10.1186/s12913-023-10331-y.

13. Dutta A, Barker C, Kallarakal A. The HIV Treatment Gap: Estimates of the Financial Resources Needed versus Available for Scale-Up of Antiretroviral Therapy in 97 Countries from 2015 to 2020. PLOS Medicine. 2015;12(11):e1001907. doi: 10.1371/journal.pmed.1001907.

14. Lahuerta M, Ue F, Hoffman S, Elul B, Kulkarni SG, Wu Y, et al. The problem of late ART initiation in Sub-Saharan Africa: a transient aspect of scale-up or a long-term phenomenon? Journal of health care for the poor and underserved. 2013;24(1):359.

15. Onoya D, Mokhele I, Sineke T, Mngoma B, Moolla A, Vujovic M, et al. Health provider perspectives on the implementation of the same-day-ART initiation policy in the Gauteng province of South Africa. Health Research Policy and Systems. 2021;19(1):2. doi: 10.1186/s12961-020-00673-y.

16. Reynolds A. Patient-centered care. Radiologic technology. 2009;81(2):133–47.

17. (NDOH) NDoHRoSA. National guidelines on conducting patient experiences of care surveys in public health establishments. Pretoria, South Africa: 2017.

18. Mokhele I, Sineke T, Vujovic M, Fox MP, Ruiter RA, Onoya D. Using intervention mapping in motivational interviewing training to improve ART uptake in Gauteng, South Africa. Journal of Health Psychology. 2022;27(3):589–600.

19. Noonan W, Moyers T. Motivational interviewing. Journal of Substance Misuse. 1997;2(1):8–16.

20. Miller WR, Rollnick S. Motivierende Gesprächsführung: Motivational Interviewing: 3. Auflage des Standardwerks in Deutsch: Lambertus-Verlag; 2015.

21. Mokhele I, Sineke T, Vujovic M, Ruiter RA, Miot J, Onoya D. Improving patient-centred counselling skills among lay healthcare workers in South Africa using the Thusa-Thuso motivational interviewing training and support program. medRxiv. 2023:2023.10. 24.23297510.

22. Schulz KF, Altman DG, Moher D. CONSORT 2010 statement: updated guidelines for reporting parallel group randomised trials. Journal of Pharmacology and pharmacotherapeutics. 2010;1(2):100–7.

23. Moyers TB, Martin T, Manuel JK, Hendrickson SM, Miller WR. Assessing competence in the use of motivational interviewing. Journal of substance abuse treatment. 2005;28(1):19–26.

24. Moyers TB, Rowell LN, Manuel JK, Ernst D, Houck JM. The motivational interviewing treatment integrity code (MITI 4): rationale, preliminary reliability and validity. Journal of substance abuse treatment. 2016;65:36–42.

25. Matenge S, Sturgiss E, Desborough J, Hall Dykgraaf S, Dut G, Kidd M. Ensuring the continuation of routine primary care during the COVID-19 pandemic: a review of the international literature. Family Practice. 2021;39(4):747–61. doi: 10.1093/fampra/cmab115.

26. Mboweni SH, Risenga PR. The Impact of The COVID-19 Pandemic on the Management of Chronic Disease in South Africa: A Systematic Review. The Open Public Health Journal. 2022;15(1).

27. Holstad MM, Essien JE, Ekong E, Higgins M, Teplinskiy I, Adewuyi MF. Motivational groups support adherence to antiretroviral therapy and use of risk reduction behaviors in HIV positive Nigerian women: a pilot study. African journal of reproductive health. 2012;16(3):14–27. Epub 2013/02/27. PubMed PMID: 23437496; PubMed Central PMCID: PMCPMC3721643.

28. Maneesriwongul W, Prajanket O-O, Saengcharnchai P. Effects of motivational interviewing or an educational video on knowledge about HIV/AIDS, health beliefs and antiretroviral medication adherence among adult Thais with HIV/AIDS. Pacific Rim International Journal of Nursing Research. 2012;16(2):124–37.

29. Zomahoun HTV, Guénette L, Grégoire J-P, Lauzier S, Lawani AM, Ferdynus C, et al. Effectiveness of motivational interviewing interventions on medication adherence in adults with chronic diseases: a systematic review and meta-analysis. International Journal of Epidemiology. 2016;46(2):589–602. doi: 10.1093/ije/dyw273.

30. Onoya D, Jinga N, Nattey C, Mongwenyana C, Mngadi S, MacLeod WB, et al. Motivational interviewing retention counseling and adherence to early infant diagnostic HIV testing schedule in South Africa: The PAEDLINK randomized trial. Medicine. 2022;101(6).

31. Gwadz M, Serrano S, Linnemayr S, Cleland CM, Cluesman SR, Freeman RM, et al. Behavioral intervention grounded in motivational interviewing and behavioral economics shows promise with Black and English-speaking Latino persons living with HIV with unsuppressed HIV viral load in New York City: A mixed methods pilot study. Frontiers in public health. 2022;10:916224. Epub 2022/10/04. doi: 10.3389/fpubh.2022.916224. PubMed PMID: 36187648; PubMed Central PMCID: PMCPMC9522600.

32. Miller SJ, Foran-Tuller K, Ledergerber J, Jandorf L. Motivational interviewing to improve health screening uptake: A systematic review. Patient Education and Counseling. 2017;100(2):190–8. doi: 10.1016/j.pec.2016.08.027.

33. Sawyer AT, McManus K. Understanding patient experiences in a motivational interviewing intervention to improve whole-person lifestyle among individuals with hypertension or type 2 diabetes: a qualitative focus group study. International journal of qualitative studies on health and well-being. 2021;16(1):1978373. Epub 2021/09/23. doi: 10.1080/17482631.2021.1978373. PubMed PMID: 34547985; PubMed Central PMCID: PMCPMC8462931.

34. Adeniyi OV, Nwogwugwu C, Ajayi AI, Lambert J. Barriers to and facilitators of HIV serostatus disclosure to sexual partners among postpartum women living with HIV in South Africa. BMC Public Health. 2021;21(1):915. doi: 10.1186/s12889-021-10955-x.

35. Tshweneagae GT, Oss VM, Mgutshini T. Disclosure of HIV status to sexual partners by people living with HIV. curationis. 2015;38(1):1–6

